# What’s On the Menu? Towards Predicting Nutritional Quality of Food Environments

**DOI:** 10.1101/2023.12.08.23299691

**Authors:** DongHyeon Seo, Abigail Horn, Andrés Abeliuk, Keith Burghardt

## Abstract

Unhealthy diets are a leading cause of major chronic diseases including obesity, diabetes, cancer, and heart disease. Food environments–the physical spaces in which people access and consume food–have the potential to profoundly impact diet and related diseases. We take a step towards better understanding the nutritional quality of food environments by developing MINT: Menu Item to NutrienT model. This model utilizes under-studied data sources on recipes and generic food items, along with state-of-the-art word embedding and deep learning methods, to predict the nutrient density of never-before-seen food items using only their name as input. The model achieves an *R*^2^ = 0.77, a substantial improvement over comparable models. We illustrate the utility of MINT by applying it to the Los Angeles restaurant food environment, and discover close agreement between predicted and ground truth nutrient density of restaurant menu items. This model represents a significant step towards a policy toolkit needed to precisely identify and target food environments characterized by poor nutritional quality.

## I. Introduction

Poor diets are a leading cause of chronic diseases such as obesity, cancer, and heart disease [1]. In 2017, an estimated 11 million deaths globally were attributed to poor dietary factors [2]. To address this issue, initiatives aimed at improving diets have been proposed by various organizations and experts in the field [3], [4]. Emerging evidence indicates that the physical environments in which individuals acquire and consume food can significantly impact diet and related health outcomes [5], [6]. Food environments impact what food is accessible and are thus recognized as a key social determinant of health [7]. Unhealthy food environments have been shown to cue unhealthy eating choices [8], [9]. Previous approaches to classify food environments have often been based on the presence or absence of broad food outlet categories [10], such as “food deserts,” environments with low supermarket access [8], [11]; and “food swamps,” environments with an over-abundance of fast food outlets [12], [13]. These broad categorizations mask the diversity of the nutritional quality of individual menu offerings (e.g., ‘fried chicken meal’ vs. ‘grilled chicken salad’); information that would support researchers and policymakers in designing interventions to increase access to healthier foods.

Our paper aims to advance the ability to assess the nutritional quality of the restaurant food environment by predicting the nutritional quality of individual restaurant menus, and aggregating across menu item predictions to come up with indicators for restaurant-level and food environment-level quality. *Nutritional quality* is a multi-faceted concept with many operational definitions [14], [15]. In this paper, we focus on a specific component of nutritional quality: *nutrient density*, the weight-based composition density of different nutrients in food items. These nutrients include both macronutrients (e.g., protein, fat, and cholesterol) and micronutrients (vitamins and minerals).

We explore this aim by starting with the hypothesis that the nutrient density of restaurant menu items can be estimated using general menu item names. This would be convenient because digital menus are relatively easy to gather. However, estimating the nutrient density of menu items from their names alone is a difficult task because menus in the real-world normally do not provide information on menu items’ nutrient composition, let alone composing ingredients or recipes. We hypothesize that nutrient density can be approximated from menu item names alone using machine learning methods. In this paper, we utilize language models to develop and evaluate methods for this task.

Towards this, we propose, introduce, and evaluate Menu Item to NutrienT (MINT), a model that predicts the nutrient density of food items from their *names* alone, by learning from large-scale meal and recipe data. We train MINT to predict a single composite index of a menu item’s overall nutrient density using the *RRR, the ratio of recommended to restricted nutrients*, an established, validated expression measuring the relationships between macro- and micro-nutrients in a food item [16]. We use multiple datasets to train and test the model. To train the model, we employ diverse food item data. This includes ‘generic’ food items, which are ‘canonical foods’ – everything from individual raw foods to complex meals – not linked to restaurant menus or recipes. We access a large dataset of generic food items providing names, nutrient composition, and ingredients from Edamam, a publicly accessible source (https://www.edamam.com/). We also use data of over one million recipes from Recipe1M+, employed to train language models that underlies a component of the prediction model. This dataset contains recipe food item names, ingredients, and preparation instructions, but not nutrient composition (for most items).

To test the model’s ability to predict the nutrient density of any kind of food item, we apply it to held-out data on generic food items from Edamam. However, because the purpose of the model is to predict the nutrition of restaurant menu items, its ultimate test of merit is in its application to restaurant menu data. It is hard to gather datasets on menus for restaurants at-large providing both the names of menu items and their nutrient composition, since as any patron will have observed, most restaurants do not publish nutrient composition information. However, the Affordable Care Act (ACA) requires that all chain restaurants with 20 or more physical outlets measure and publish nutrient information on each of their menu items [17]. A dataset on chain restaurant menus and nutrient information is compiled by Spoonacular, and made publicly available via their API (https://spoonacular.com/food-api). We source this data to provide an evaluation of our model on real-world, out-of-sample restaurant menu data. The Spoonacular dataset provides food items gathered from real chain restaurant menus, such as McDonald’s, Subway, Sweetgreen, and Starbucks.

We evaluate the model’s ability to predict nutrient density at three levels of analysis – individual menu items, restaurant menus, and across all restaurants in our dataset in a neighborhood food environment. To address the latter, we link the Spoonacular dataset to an additional database providing the location of restaurants in LA County.

To summarize, our contributions are as follows:

1) We develop a state-of-the-art open-source method to predict the nutrient density of restaurant menu items ^1^.
2) We perform a qualitative evaluation of the model using several ground truth datasets, demonstrating strong predictive performance as well as substantive improvements against baselines and state-of-the-art algorithms.
3) We use the model to evaluate the Los Angeles chain restaurant food environment using large-scale, realworld Chain restaurant menu data and find good agreement with ground truth data.

These contributions are a substantial step towards a fine-grain evaluation of food environments, a critical toolkit for policy-makers aiming to improve the diets and health of the general population by identifying and targeting areas characterized by poor nutritional quality.

## II. Related Works

### A. Food item nutrient density indicators

The leading cause of dietary disease is the under-consumption of recommended nutrients and food groups [18], rather than the overconsumption of calories, as nutrient distribution and caloric value have been shown to be largely uncorrelated within food items [19], [20]. Towards this purpose, several continuous indicators have been designed for evaluating the composition of nutritional components in a diet or food item [21], [22]. These include the Healthy Eating Index (HEI) [23], which assesses amounts of key dietary components (i.e., total fruit, whole fruit, legumes, etc.), and several indices assessing *nutrient density*.

Nutrient density scoring of individual food items has been demonstrated to be a reliable and valid tool for quantifying nutritional quality in a single score [16], [24], [25]. Multiple regulatory applications have employed these tools including evaluating labeling and marketing of snack foods, food tax programs, and defining school food standards [25]–[These include the Ratio of Recommended to Restricted nutrients (*RRR*) [16], used in this paper, and the Nutrient Rich Foods Index (*NRF*) [24], [31].

The *RRR* [16] is the average of the % of the FDA recommended daily value (RDV) of each recommended nutrient, namely protein and fiber (macronutrients), as well as vitamins A and C and minerals Calcium and Iron (micronutrients), divided by the average of % of the RDV for restricted nutrients, namely sugar, saturated fat and calories (macronutrients) and sodium and cholesterol (micronutrients):

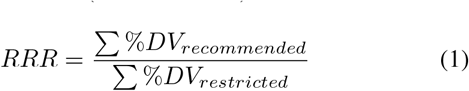

Here, we show the daily value intake for each nutrient that makes up *RRR* in Table I; they are defined and recently updated by FDA [32]. We use this value when calculating the nutrient density scores.

**TABLE I.** Daily value Intake for each nutrient consists of *RRR*.

| Nutrients | Daily Value Intake |
| --- | --- |
| Protein | 50(g) |
| Fiber | 28(g) |
| Vitamin A | 900(μg) |
| Vitamin C | 90(mg) |
| Calcium | 1300(mg) |
| Iron | 18(mg) |
| Calorie | 2000(kcal) |
| Total Sugar | 50(g) |
| Cholesterol | 300(mg) |
| Saturated Fat | 20(g) |
| Sodium | 2300(mg) |

### B. Food computing

A range of recent literature has evaluated how to extract properties of food from images and text [33]. This includes predict recipe [34] or nutrition information from food images [35]–[37], but also a proliferation of methods to match food names to recipes and macro-nutrients [33].

Many of these methods (e.g., [37]) are alike to Liu et al., which developed an algorithm to estimate the nutritional quality of food items by matching names to food items in the USDA National Nutrient Database for Standard Reference (NNDSR), the lab-verified gold-standard of the nutrition composition of foods in the U.S. that contains around 8,000 generic and branded food items [38], [39]. To find matches to the NNDSR, the *USDA matching method* [38] makes use of the algorithm made available through the FoodData Central API, which is based on ElasticSearch, a character-based matching algorithm for computing a similarity measure between two strings. While these and other methods also match food names to macronutrients [33], menus often contain food names that do not appear in these datasets, motivating the need for the new methodology we develop in this paper.

### C. Text embedding

Word embeddings, such as those used to extract food properties from text [33], have a long history, including Word2Vec [40], an improvement over even earlier Bag-of-word vectors [41]. FastText was an improvement on these previous methods due to its ability to map words, even words not seen in the training set, to embeddings. Alternatives and improvements to this model include GLoVE [42], and transformer-based contextual embeddings, most notably BERT [43] and RoBERTa [44]. More nuanced embedding methods have been proposed, including those that embed entire sentences, such as Sentence-BERT (SBERT) [45], or longer documents, such as Longformer [46]. The predictive performance of models using text embeddings is domain-dependent, motivating the need to test different text embedding methods in new applications.

### D. Clustering and Fine-tuning

Many clustering methods exist, most notably K-Means [50], which partitions data to their nearest cluster centroid. The number of clusters is determined from metrics, such as the elbow method [51], or Silhouette score [52]. An alternative approach to this method is DBSCAN [53], in which the authors defined clusters based on the density of points, and developed a way to determine the number of clusters. Both of these methods, however, create “hard” clusters while most data are unlikely to partition so cleanly into a given cluster. We therefore use HDBSCAN, a method similar to DBSCAN that efficiently performs soft clustering, which allows for data to belong to multiple overlapping clusters. Further variations of these ideas exist (cf. [54]).

Fine-tuning used in this paper is a common approach to improving models with limited data and is often used in AI tasks [55], [56], including NLP tasks [57], and multi-label classification tasks [58]. These methods address a common problem of *distribution shift* [59], where the distribution of features varies across domains, and *covariate shift* [60], [61], *where the relationship between features and outcome variables differs across domains*.

### E. Our contribution

This paper improves upon the previous methods by leveraging state-of-the-art word embedding models and feed-forward neural networks to interpolate the nutritional score of menu items. We aim to train the model to infer the nutrient values of food items without the need for precise ingredient identification using clustering to get food category pseudo-labels and fine-tuning techniques. The approach is possible due to the large scale of generic food items in our training data, as described below.

## III. Methods

We aim to develop MINT to predict the nutrient density of individual restaurant menu items and, ultimately, an overall nutrient density score for each restaurant. We use the nutrient density indicator *RRR* [16], as defined in Equation 1. We choose *RRR* over the other approaches and indices for evaluating the nutritional components of food items because (i) our validation data provides access to nutrient values rather than food groups, making the use of recommended nutrients more straightforward than approaches assessing key dietary components (i.e. food groups); and (ii) we do not attempt to predict the portion size of a menu item in this work; this allows us to use the *RRR*, which is portion-size invariant due to its ratio construction, but not the *NRF*.

We also created a similar metric *RRR*_*macro*_, which is *RRR* without micronutrients in the numerator (*RRR*_*macro*_). *RRR*_*macro*_ was used where micronutrients were inaccurate or missing in datasets (i.e. Spoonacular).

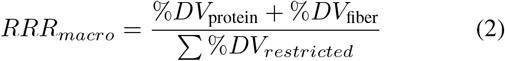

The pipeline for the MINT framework is shown in Fig. 1. This algorithm extracts a nutrient density score from each menu item in three steps. We first utilize a range of data to create specialized language models for extracting features [48]. Next, we use a state-of-the-art sentence embedding model, MPNet [49] to embed menu item names with their ingredients to create ingredient-contextualized food clusters. These clusters act as food category pseudo-labels to train a model that maps menu item names (without their ingredients) to the learned pseudo-labels. Finally, we train a model that predicts menu item nutrient density and then fine-tune copies of that model to each food category pseudo-label.

**Fig. 1.**
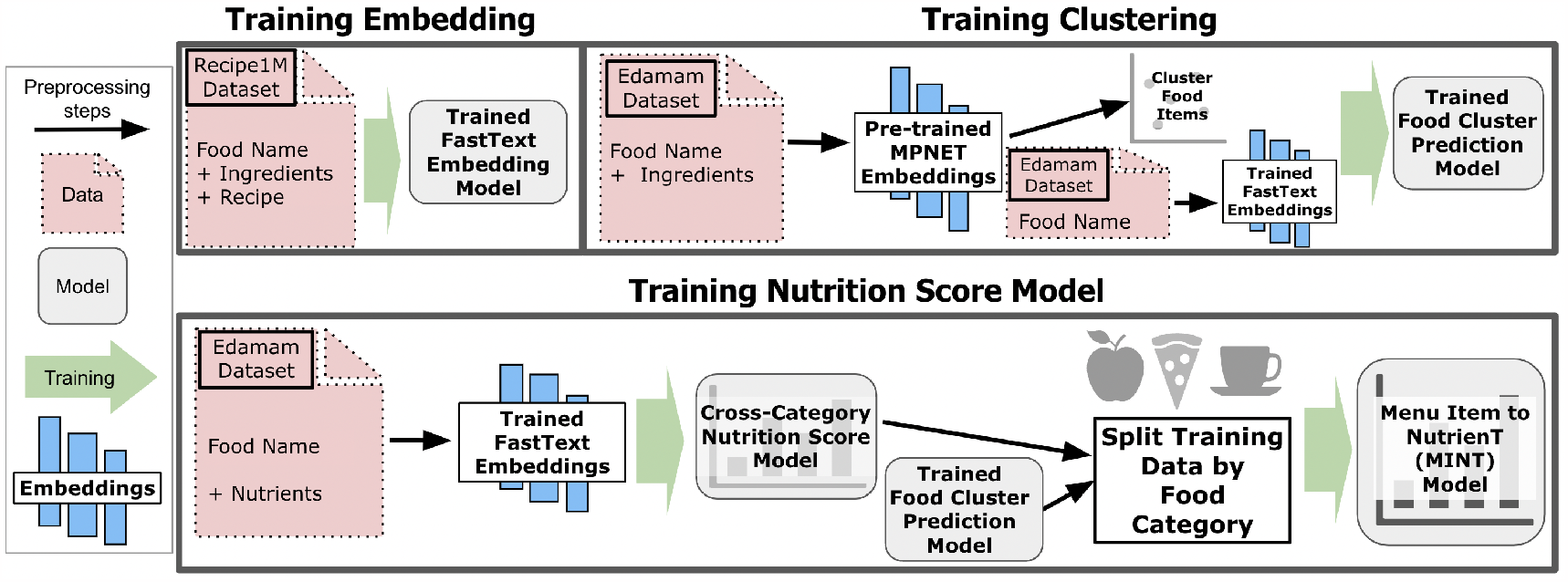
Nutrition prediction pipeline. (a) **Word embedding model**. We begin by extracting food names, ingredients, and recipes from the Recipe1M+ [34], [47], and use the concatenation of this text to train a FastText word embedding model [48]. (b) **Food category prediction model**. We embed Edamam training data containing menu items concatenated with their ingredients using a pre-trained MPNet model [49]. We then cluster the training data using HDBSCAN, which we treat as a ground truth food category. We use the Edamam FastText embeddings from food names alone to train a model to predict the most likely food category associated with each food name. (c) **Nutrition Score Model**. The FastText embeddings are used to train a model to predict the nutrient density score. The model first trained on the entire dataset is then fine-tuned on the ground-truth categories. Finally, MINT predicts food item nutrient density conditional on the predicted food category.

### Data

#### 1) Edamam

We extract food items, ingredients, and nutrients from the publicly available Edamam Food Database API ^2^ after being granted permission from the company to store these data. The dataset consists of 81,390 generic food items, which is based on recipe and food composition data from multiple sources, including the U.S. Department of Agriculture’s (USDA) Food Data Central [62], and curated to ensure the accuracy of these data. A total of 33 different macro- and micronutrients were available. We use a subset of these that are most complete: Protein, Calories, Fiber, Total Sugar, Vitamins A & C, Cholesterol, Saturated Fat, Calcium, Sodium, and Iron.

#### 2) Spoonacular

We also extracted a separate food item nutrient density dataset, Spoonacular, via its API ^3^. This dataset provides the names and nutrient composition of menu items from chain restaurants, defined as restaurant brands with 20 or more physical outlets [17]. We also extracted menu items from 9 additional chain restaurant brands, which were not in Spoonacular’s database but are prevalent within the Los Angeles County area - their menus and nutrition information were manually gathered from each of their official websites. The specific restaurants we manually curated were: The Cheesecake Factory, Chipotle, El Torito, Flame Broiler, Hot Dog On A Stick, Mendocino Farms, Pick Up Stix, Raising Canes, and Tender Greens.

In total, these data include 24K menu items across 145 large restaurant chain brands with 4663 physical outlets in LA County, but contained incomplete and sparse micronutrient values. We use these data as an out-of-sample test of MINT’s performance in predicting nutrient density at three levels of analysis: individual menu items, restaurant menus, and across all restaurants in our dataset in a neighborhood.

#### 3) Recipe1M+

We also utilized Recipe1M+ [34], [47], a large-scale structured dataset containing over 1 million cooking recipe triplets, which includes food item names, ingredients, and nutrient values, as well as instructions for preparation (recipes). The nutrient values are too incomplete to use in our analysis and are therefore removed. The other features are used to train a FastText model and fine-tune BERT model.

### B. Transferring Ingredients Information with Food Category Pseudo-Label

Most restaurant menus only provide a name for items without a description of ingredients or preparation. Predicting nutrients based just on a name is a challenging task. However, predicting the nutrients may be easier if we know what kind of food the item is, e.g., a dessert versus a salad. We take a page from AI Fairness to address this issue [63], and build MINT to first predict a broad food item category by using pseudo-labels created from the clustering task, and then predict the nutrients from a model fine-tuned to that category. This categorization step improves our predictions compared to a single one-size-fits-all model trained on the entire dataset across all categories.

To group the food items, we take **both** *food item names* and *ingredients* in the Edamam dataset by converting these data into a sentence “*Food Name* made with *query of ingredients*”, for example, “*Egg scrambled* made with *butter, egg, salt, pepper*.” We then embed these texts using a pre-trained MPNET model [49], map it to a lower dimension space, and finally cluster these embeddings to create food category pseudo-labels for each food item.

We use UMAP [64] to reduce the dimension of embeddings to two, minimizing the curse of dimensionality (where embeddings perform poorly in high dimensions [65]). We then cluster using HDBSCAN [66], which allows for items to belong to multiple clusters (e.g., a chicken burger could be a chicken dish or burger). Hyperparameters, such as dimension (2,10,50) or “*n neighbors*” (10,20,50) for UMAP, and “*min samples*” (10, 20, 50), “*min cluster size*” (1000), as well as cluster selection epsilon (0.0, 0.05, 0.10) from HDBSCAN do not significantly affect results.

An example of the clusters is shown in Fig. 2a, and the distribution of the nutrient density scores is shown in Fig. 2b and 2c. We notice clear differences in both nutrient density scores, *RRR* and *RRR*_*macro*_, between clusters.

**Fig. 2.**
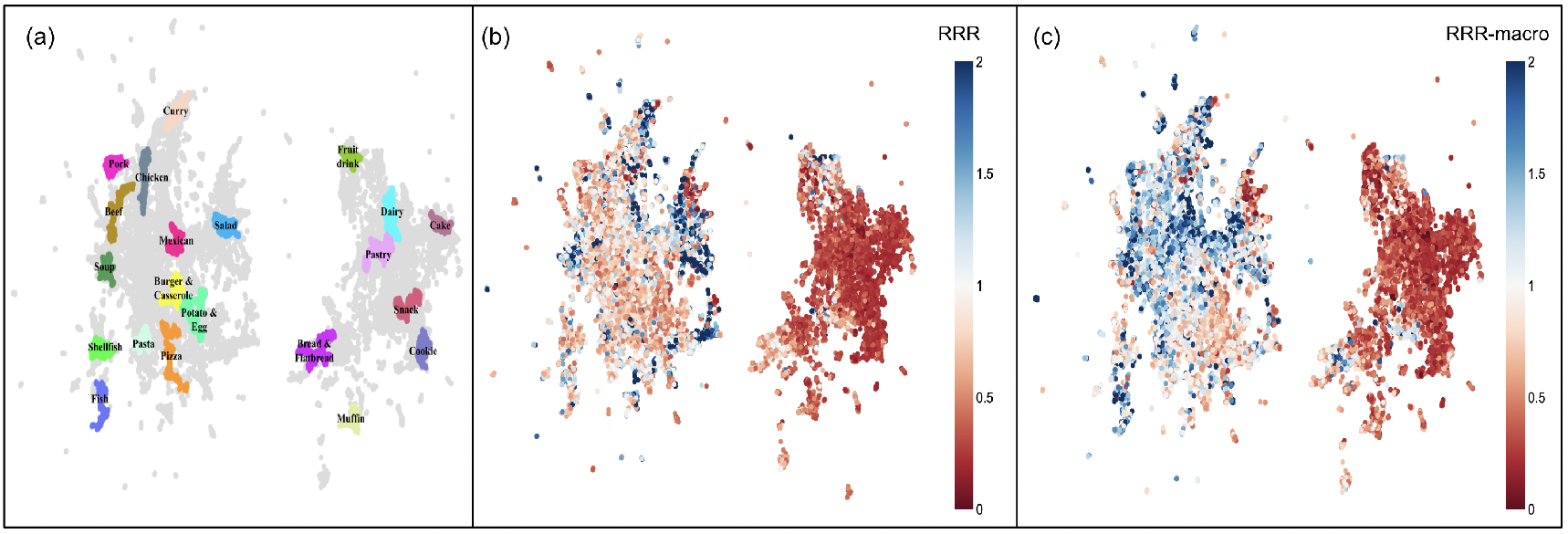
Food item and ingredient embeddings. (a) Each color represents high-confidence exemplars of each food item cluster label. (b) The nutrient density (*RRR*_*macro*_) for food items on the embedding map. (c) The nutrient density (*RRR*) for food items on the embedding map.

### C. Creating Food Language Vector Space

Next, we embed food items both to predict clusters and to predict nutrient density scores. We train a FastText model [48] using the Recipe1M+ dataset (**RecipeFT**) [34], [47], which contains 1.02M entries of food names, ingredients, and recipes (∼ 171M tokens in total), to construct a vector space tailored to language in the food domain. We create embeddings by averaging vectors of all the words in each food name. All of the embeddings have a 300-dimensional latent space. We also fine-tune the masked language model of BERT [43] using the Recipe1M+ (**RecipeBERT**) to compare these two different approaches to language modeling. We extract features by adding a mean pooling layer to take the attention mask into account and get embeddings with 768-dimensional space.In our ablation study, we compare MINT’s Recipe1M+ dataset-based embeddings to respective pre-trained FastText [48] and BERT [43] models. Model weights for RecipeFT and the repository for RecipeBERT are included in the shared repository^1^.

### D. Predicting Nutrient Density of the Food Item

#### Category pseudo-label prediction

Recall MINT requires predicting food item categories and linking food items to their category-specific models. To create a ground truth food category for each food item, we take their most likely cluster to be their category pseudo-label. To predict the category of food item names, we train a five-layer Multi-Layer Perceptron (MLP) where inputs are word representations of each food item name, and their outputs are predicted categories. We train this model using binary cross entropy loss and softmax activation in the final layer with early stopping using a 20% of training dataset as validation set [67], [68]. Numbers of hyperparameters [69]–[74] were tested for both the category and nutrition prediction models that use MLP, shown in Table II.

**TABLE II.** Hyperparameters tested for MLP.

| Hyperparameter | Value |
| --- | --- |
| Number of Nodes in Each Layer | [128, 256, 512] |
| Activation Function | [*ReLU*, *LeakyReLU*($\alpha = 0.01$), *LeakyReLU*($\alpha = 0.1$)] |
| Dropout Rate | [0.1, 0.2, 0.5] |
| L2 Regularization $\lambda$ | [$1e-2$, $1e-3$, $1e-4$] |
| Learning Rate | [$1e-2$, $1e-3$, $1e-4$] |
| Input Normalization | [*True*, *False*] |
| Layer Normalization | [*True*, *False*] |
| Batch Normalization | [*True*, *False*] |

#### Food Category Weighted Mean Baseline

We use the category prediction model to create a food category weighted mean (FCWM) baseline nutrition density model. Because Fig. 2b and 2c show substantial differences in nutrient density scores between categories, we expect this to be a strong baseline. We first find the confidence, *p*_*i*_, a food item has in each food category, *i*. Then, multiply them with the true mean (TM) of the nutrient density score of their labels. FCWM is

then:

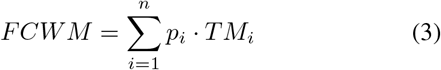

#### ML Baseline

We contrast this FCWM baseline method with a nutrient density predicting Machine Learning (ML) model. We set the end-to-end pipeline from language model to prediction model without additional structure. We explored few state-of-the-art ML prediction models including *Ridge Regression* [75], *Random Forest* [76], *xgboost* [77], and *MLP*, while MLP retained the best results among different models. Therefore, we use MLP for our ML baseline prediction method. Similar to the category prediction model, we train on a five-layer MLP where the input is a food item name embedding, and the output is a *RRR* or *RRR*_*macro*_. We train this model by minimizing the mean squared error between predicted and actual nutrient density and using a linear activation function in the final layer with early stopping using a 20% of training dataset as validation set [68], [78].

#### MINT

Finally, we combine category prediction and the MLP method to create MINT. After training the MLP model, we split the training dataset by the ground truth food categories and fine-tune copies of this model for each food category. This approach of utilizing the pseudo-labels for unlabeled data has shown great improvement in various supervised learning tasks by posing them in a semi-supervised learning fashion [79]. While training the category-specific models We keep the Adam learning rate the same but add weight decay set at 0.001, to avoid local minima [80]. Although each model is fine-tuned using ground truth food category pseudo-labels, we use the predicted food categories on the held-out data as they do not have true food category labels.

For both the MLP and MINT methods, we also utilize MC Dropout [71] to create error estimates of predictions with 100 MC iterations and produce 95% confidence intervals. We show examples of predictions with confidence intervals in Table III.

**TABLE III.** Prediction examples using MINT.

| Food Item | True $RRR$ | Predicted $RRR$ (95% CI) |
| --- | --- | --- |
| Poutine | 0.845 | [0.838, 0.849] |
| Ratatouille Provencale | 1.557 | [1.544, 1.562] |
| Cinnamon Swirl Rolls | 0.271 | [0.269, 0.273] |

### E. Restaurant Nutrient Density Score

As our goal is to measure the nutrient density of restaurant menus overall rather than individual food items, we also introduce *Restaurant Nutrient Density* (*RND*), defined as the median of restaurant menu items’ *RRR*_*macro*_. We specifically use *RRR*_*macro*_ for this case, as the Spoonacular dataset with Restaurant menu items does not contain some of the micronutrients included in *RRR*. We measure and qualitatively compare *RND* in the Los Angeles chain restaurant environment. To extract the locations of each restaurant, we use the Los Angeles County Restaurant and Market Inventory [81], which contains the locations for all registered restaurants in Los Angeles County. We use these locations to evaluate the spatial distribution of chain restaurants *RND* across Los Angeles County. Individual menu items with outlier *RRR*_*macro*_ values were removed from the ground truth data using an outlier detection method, histogram-based outlier score [82], thus reducing *RRR*_*macro*_’s range from well over 100 to 2.82 while removing just 762 menu items out of 24748 (3% in total). Results are qualitatively the same if we include these data.

## IV. Results

In this section, we show the performance of MINT, including its performance over the previous state-of-the-art and against ablated models.

### A. Ablation Study

We created an ablation study of our model, MINT on Table IV for both *RRR*_*macro*_ and *RRR*. The best-performing model and text embedding data, which we use throughout the study, are in bold. To ablate this model, we removed food category prediction and fine-tuning from MINT, leaving us with an end-to-end pipeline with ML prediction head (MLP). Next, we removed a nutrient prediction head while using food category prediction to approximate nutrient density score from the weighted mean (FCWM), and compared these with MINT. We test these models using embeddings derived from pre-trained BERT (base-uncased), BERT fine-tuned with the Recipe1M+ dataset, and FastText embeddings trained on Common Crawl, and text from the Recipe1M+ dataset.

**TABLE IV.** Competing nutrient density scores prediction models.

| NLP Model | Prediction Model | $R^2 \uparrow$ | | $RMSE \downarrow$ | | $MAE \downarrow$ | |
| --- | --- | --- | --- | --- | --- | --- | --- |
| | | $RRR$ | $RRR_{macro}$ | $RRR$ | $RRR_{macro}$ | $RRR$ | $RRR_{macro}$ |
| Pre-trained (base-uncased) BERT | FCWM | 0.267±0.002 | 0.363±0.013 | 0.281±0.001 | 0.241±0.002 | 0.332±0.005 | 0.338±0.005 |
|  | MLP | 0.591±0.019 | 0.658±0.01 | 0.159±0.006 | 0.129±0.005 | 0.227±0.004 | 0.233±0.004 |
|  | MINT | 0.624±0.009 | 0.689±0.01 | 0.145±0.006 | 0.120±0.004 | 0.216±0.003 | 0.219±0.002 |
| RecipeBERT | FCWM | 0.296±0.006 | 0.399±0.017 | 0.273±0.005 | 0.227±0.008 | 0.324±0.006 | 0.326±0.007 |
|  | MLP | 0.633±0.009 | 0.705±0.007 | 0.142±0.005 | 0.112±0.004 | 0.211±0.006 | 0.214±0.002 |
|  | MINT | 0.667±0.008 | 0.729±0.008 | 0.129±0.003 | 0.103±0.004 | 0.197±0.003 | 0.200±0.003 |
| Pre-trained (Common Crawl) FastText | FCWM | 0.301±0.008 | 0.405±0.02 | 0.270±0.002 | 0.226±0.001 | 0.323±0.004 | 0.326±0.004 |
|  | MLP | 0.671±0.013 | 0.732±0.01 | 0.128±0.006 | 0.102±0.004 | 0.203±0.005 | 0.198±0.002 |
|  | MINT | 0.688±0.014 | 0.757±0.007 | 0.121±0.005 | 0.093±0.004 | 0.190±0.002 | 0.189±0.002 |
| RecipeFT | FCWM | 0.300±0.007 | 0.406±0.019 | 0.270±0.002 | 0.226±0.001 | 0.322±0.005 | 0.325±0.004 |
|  | MLP | 0.691±0.01 | 0.750±0.008 | 0.120±0.004 | 0.099±0.004 | 0.194±0.002 | 0.195±0.001 |
|  | MINT | **0.710 ± 0.01** | **0.768 ± 0.006** | **0.111 ± 0.003** | **0.088 ± 0.003** | **0.182 ± 0.002** | **0.183 ± 0.001** |
| USDA-matching algorithm [38] |  | -9.82±2.72 | -2.49±0.745 | 4.376±0.957 | 1.405±0.314 | 0.637±0.018 | 0.569±0.01 |

We use one-way ANOVA [83] and Tukey’s range test [84] to determine if there is a significant difference in *R*^2^ across the various ablation methods. We find that MINT outperforms all competing methods with statistically significant ANOVA p-values *<* 10^*−*10^. The different FastText word embeddings are borderline significant with ANOVA p-value = 0.047. BERT and RecipeBERT perform worse indicating a FastText, despite its relative age, can be more useful for MINT.

### B. Validation Study

We compare our method against the current state-of-the-art, the *USDA matching method* [38]. The results on the Edamam held-out dataset, shown in Table IV and Fig. 3, show that MINT performs substantially better for both nutrient density metrics. Using *RRR*_*macro*_, MINT performs prediction with *R*^2^ = 0.77, compared to *R*^2^ = −2.5 for the USDA matching method. An *R*^2^ *<* 0 implies that the results are worse than guessing the average *RRR*_*macro*_. The RMSE and MAE performance metrics show similar behavior. In addition, the USDA matching method can only match 53% of held-out data compared to MINT, which calculates 100% of menu items. The results are even worse for the *RRR* metric that includes micronutrients, where only 27% are matched using the USDA matching method.

**Fig. 3.**
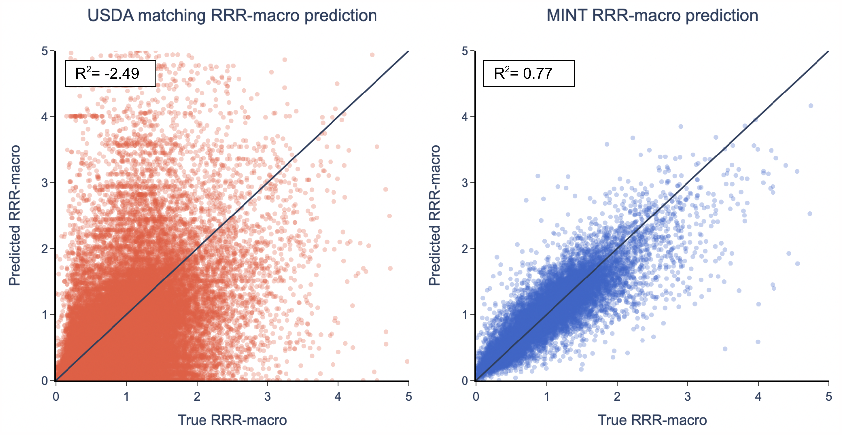
Predicting nutrient density (*RRR*_*macro*_) against state-of-the-art algorithm. Results are for 20% held out data from Edamam. (a) The USDA-matched algorithm [38] (*R*^2^ =− 2.49, 53% of data matched). (b) MINT (*R*^2^ = 100.77; 100% of data matched).

We tested the generality of MINT by predicting the nutrient density of individual food items in the Spoonacular dataset when MINT was trained on Edamam. As we show in Fig. 4, MINT has a performance of *R*^2^ = 0.31, while the USDA matching method has *R*^2^ = − 2.6, which indicates the model again performs worse than naively guessing the mean *RRR*_*macro*_. Similar to the Edamam dataset, the RMSE and MAE metrics show similar behavior as shown in Table V. Finally, the USDA matching method only finds the nutrient density for 67% of food items.

**Fig. 4.**
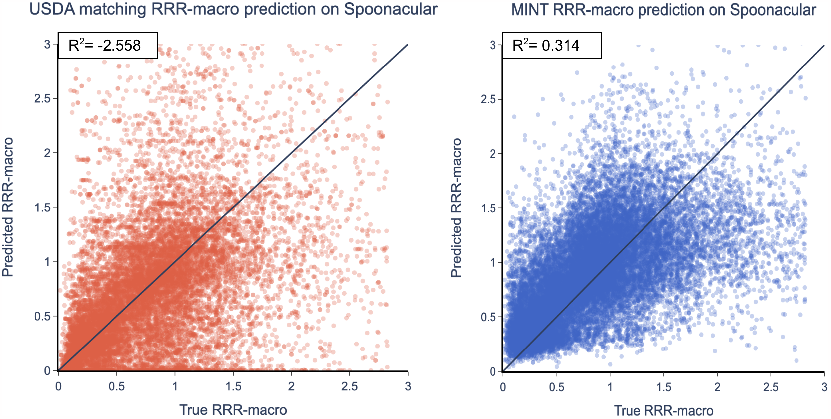
Predicting nutrient density (*RRR*_*macro*_) against state-of-the-art algorithm. The results are for the Spoonacular dataset. (a) The USDA-matched algorithm [38] (*R*^2^ = − 2.56, 67% of data matched). (b) MINT (*R*^2^ = 0.31; 100% of data matched).

### C. Restaurant Nutrient Density

We demonstrate the performance of MINT to predict restaurant nutrient density on a large-scale real-world restaurant database in Fig. 5. This figure shows the *RND* averaged across all chain restaurants in Los Angeles County neighborhoods [85] using Fig. 5a MINT and Fig. 5b ground truth data. We find (i) that *RND* of chain restaurants varies substantially across neighborhoods within Los Angeles County, and (ii) that MINT predictions closely match ground truth *RND*. Overall, MINT has an *R*^2^ = 0.46 between the predicted and ground truth *RND* across 145 restaurant chains, while the USDA matching method has an *R*^2^ = 0.27, as shown in Table V. This result points to two critical advantages of MINT: first, menu items and *RND* scores can be inferred despite the lack of standard approaches to naming menu items across restaurants. Second, while simple matching is not powerful enough for our needs [38], the MINT predictive model can generalize well to out-of-sample data.

**TABLE V.** Model performances on individual menu item *RRR*_*macro*_ AND MEDIAN *RRR*_*macro*_ BY EACH RESTAURANT’S MENU: Restaurant Nutrient Density (*RND*) using Spoonacular dataset.

| Prediction Model | Metrics | $R^2 \uparrow$ | $RMSE \downarrow$ | $MAE \downarrow$ |
| --- | --- | --- | --- | --- |
| USDA-matching algorithm [38] | $RRR_{macro}$ | -2.56±1.48 | 1.019±0.426 | 0.459±0.014 |
| MINT | $RRR_{macro}$ | **0.314 ± 0.011** | **0.200 ± 0.006** | **0.322 ± 0.004** |
| USDA-matching algorithm [38] | $RND$ | 0.271±0.224 | 0.063±0.009 | **0.175 ± 0.02** |
| MINT | $RND$ | **0.457 ± 0.099** | **0.059 ± 0.011** | 0.180±0.018 |

**Fig. 5.**
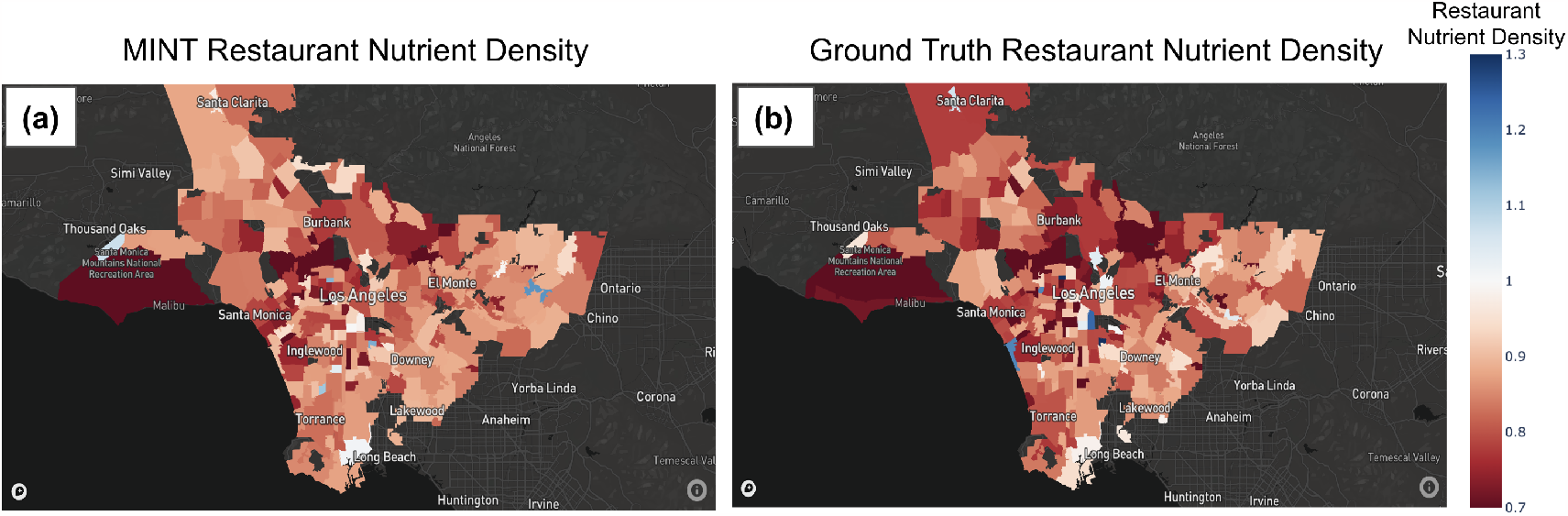
Restaurant nutritional quality in Los Angeles County. These plots show the median *RRR*_*macro*_ for each restaurant menu, or *RND*, averaged across all chain restaurants in neighborhoods. (a) MINT restaurant quality predictions and (b) Ground truth restaurant quality.

## V. Discussion

Our results demonstrate that MINT is a novel method to extract nutritional information from menu items without text matching. We find that it greatly outperforms competing methods, with *R*^2^ = 0.77 for the nutrient density score, *RRR*_*macro*_, in held-out data. Because our method predicts rather than matches the text, we can capture the nutrient density scores for 100% of food items rather than 53% for the previous state-of-the-art [38]. Additionally, MINT provides the pseudo-labels of the type of food each menu item was classified to (e.g., Chicken dish, Salad, Cake, etc.).

Having demonstrated the utility of MINT, we revisit and evaluate the aims we posed for this paper.

### • Menus Can Measure Restaurant Nutrient Density (*RND*)

We show that menus are easy to gather, and that nutrient density scores can be predicted from their composing items, *RRR* and *RRR*_*macro*_. This, in turn, allows us to accurately estimate *RND*, the average nutrient density scores across menu items. *RND* acts as a good proxy of nutritional quality as we qualitatively find healthier restaurants (e.g., restaurants serving salad-based meal items such as Sweetgreen) have high *RND*, while unhealthy restaurants (such as restaurants serving mostly low-nutrient energy dense foods such as McDonald’s), have low *RND*. Despite not knowing restaurants’ portion sizes, the median nutrient density across all items in a menu allows for a strong estimate of the nutrient density across a restaurant, RND.

### • Food Item Names Can Be Sufficient to Determine Nutrient Density

We hypothesized that we could approximate the nutrient density of the food item using only their names as input. The performance of MINT provides compelling evidence that the nutrient density of menu items can be estimated using publicly available menu data using ML prediction techniques. A key strength of the model is the introduction of the fine-tuning of models to groups with similar ingredients. This allows food items like “Buddha’s Delight” which have non-conventional food names that provide no direct information on composing ingredients, to be grouped with items with similar ingredient profiles, preventing MINT from miscalculating the nutrient density scores through covariate shifts and out-of-distribution instances [86], [87]. Overall, we find that food item names accurately represent the nutrient density of food items. Moreover, NLP techniques, such as creating food language vector space using language models like Fastext or building domain-specific Large Language Model (LLM), allow for nutrient density to be more accurately calculated than competing methods.

### • Predicting Nutrient Density With Public Datasets

We demonstrate that three different publicly available datasets used for different purposes (Edamam–training prediction models; Spoonacular–validation on out-of-sample, realworld restaurant data; Recipe 1M+ [34]–to construct food language vector space for text embeddings) allow development of a model applicable to both food item and restaurant nutritional quality scores.

## VI. Conclusions

Public health nutrition research seeks to evaluate the impact of access to (un)healthy food environments on diet quality and health outcomes. Commonly used food environment indicators are based on the presence of food outlet categories. This study, in contrast, introduces a state-of-the-art deep learning-based approach to predict nutrient density, a component of nutritional quality, of restaurant menu offerings. Strong prediction results on out-of-sample data demonstrate that MINT can robustly impute nutrient density values for the many restaurant menus that do not measure or publish their nutritional composition information. This algorithm is a significant step to help policymakers assess nutritional disparities.

There are several key limitations to our method warranting future work. With regard to the datasets used for validation, we evaluated MINT using out-of-sample restaurant data from large chain restaurants only, meaning our results may not generalize to non-chain restaurants. However, the chain restaurants in the Spoonacular dataset represent a distribution of cuisines and food types served (e.g., cafes like Starbucks to traditional fast food outlets like McDonald’s to modern chains focusing on salad, Sweetgreen), and demonstrate a wide distribution of nutrient density based on ground truth *RRR*_*macro*_. Additionally, the Edamam dataset we trained and evaluated our model on contained diverse ‘generic food items’ that are not specific to chain restaurants. We therefore believe that this limitation will not significantly affect our results.

Additional limitations are particular to the measures of restaurant and food environment nutrient density we define. First, the data we train on does not include portion sizes; therefore, MINT cannot determine whether the unhealthy or healthy menu items are a large or small contribution to someone’s overall food intake. Moreover, while our method focuses on the nutrient density of restaurant menus, there are other dimensions of restaurant nutritional quality as well. This might include the foods most commonly ordered, food safety, freshness, taste, price, and appearance. Future work should infer nutritional quality from these and other dimensions, especially with the help of multi-modal (e.g., text and image) analysis.

## Data Availability

The data used in this study were accessed through public domain websites and APIs. The Edamam data on generic food items is available from the Edamam Food Database API, found at: https://www.edamam.com/. The Spoonacular data on chain restaurant menus is available from their API, found at https://spoonacular.com/food-api. The Recipe1M+ data can be accessed directly from their website at http://pic2recipe.csail.mit.edu/.

https://github.com/alexdseo/mint

## Acknowledgments

This work was funded by the National Institute of Minority Health and Health Disparities (NIMHD) of the NIH under award number P50MD017344, to the Southern California Center for Latino Health (SCCLH).

We would like to thank Edamam for sharing and allowing us to locally store their curated generic meal item data.

We would also like to thank May Wang, Professor of Community Health Sciences at UCLA, for advising discussions regarding the restaurant and food environment level indicators.

## Footnotes

1 Our code is available here: https://anonymous.4open.science/r/mint-B5F3.

2 https://www.edamam.com/

3 https://spoonacular.com/food-api

